# Germline HLA-DQ genotype influences response to CAR T-cell therapy in patients with large B-cell lymphoma

**DOI:** 10.1101/2024.01.11.24301087

**Authors:** Melchior Le Mene, Vincent Allain, Juliette Villemonteix, Alexis Cuffel, Jean-Luc Taupin, Roberta di Blasi, Catherine Thieblemont, Sophie Caillat-Zucman

## Abstract

CD19-directed chimeric antigen receptor (CAR) T cells have greatly improved the prognosis of relapsed/refractory large B-cell lymphoma (rrLBCL), yet treatment failure occurs in more than half of patients, usually in the first 3 months after treatment. While they primarily act through CAR-dependent, HLA-independent recognition of tumor targets, CAR-T cells may also indirectly contribute to long-term tumor immunosurveillance by stimulating endogenous immunity. We hypothesized that HLA diversity, measured by the HLA evolutionary divergence (HED) metric which reflects the breadth of the immunopeptidome presented to host T cells, could influence antitumor response after CAR T-cell therapy, as seen after immune chekpoint inhibitor treatment. We studied 127 rrLBCL patients treated with commercial CAR-T cells in our center, of whom 50 % achieved durable response. We observed no impact of diversity at any HLA locus, except for HED-DQA1 that was surprisingly negatively associated with response. Analysis of the distribution of HLA-DQ alleles according to clustering of HED values pointed to the DQ1/DQ1 genotype as an independent predictor of durable response and lower incidence of relapse/progression. These findings highlight the unsuspected role of germline HLA-DQ molecules in the response to CAR-T cells and suggest an important contribution of cross-talk between CAR-T cells and endogenous immune cells.

**Key Points:**

- Germline HLA-DQ genotype is an independent predictor of durable response and lower incidence of relapse/progression after CAR T-cell therapy in rrLBCL
- HLA-DQ1/DQ1 genotype could influence the host immune response after CAR T-cell therapy and increase the chances of a durable response

## Introduction

Chimeric antigen receptor (CAR)-T cells targeting CD19 have revolutionized the treatment of relapsed/refractory large B-cell lymphomas (rrLBCL), yet less than half of patients show durable response. Among factors associated with CAR T-cell treatment failure, the emergence of CD19-negative escape variants and lack of CAR T-cell persistence are important causes of tumor relapse^1,2^. In addition to their HLA-independent, on-target recognition of CD19^+^ tumor cells, CAR-T cells can boost the host immune response, which helps limit antigen-loss-mediated relapse and likely contributes to long-term tumor immunosurveillance^3,4^.

The highly polymorphic HLA class 1 and class 2 molecules bind and present antigenic peptides to CD8 and CD4 T lymphocytes respectively, subsequently initiating antigen-specific immune responses. According to the heterozygote advantage^5–7^, heterozygous HLA alleles present a broader repertoire of antigenic peptides to T cells, which in turn promotes a more diverse T-cell response. HLA diversity can be further quantified by the HLA evolutionary divergence (HED) metric, which reflects the breadth of the immunopeptidome presented to T cells^8^ and in turn, the strength of immune response. HED is associated with the response to immune checkpoint inhibitors in cancer patients^9–11^ and with the outcomes of hematopoietic stem cell or solid organ transplantation^12–15^. Notably, HLA heterozygosity and higher HED have recently been associated with a reduced risk of cancer development, including non-Hodgkin lymphomas^16,17^.

Herein, we hypothesized that HLA diversity, by increasing the repertoire of tumor antigens presented to endogenous T cells, could promote the host immune response after CAR T-cell therapy and increase the chances of a durable response.

## Methods

We performed a retrospective analysis of all patients who received commercial CAR-T cells for rrLBCL at St-Louis Hospital between June 2018 and January 2022 and had available DNA sample for HLA genotyping. Patients received axicabtagene-ciloleucel (axi-cel) or tisagenlecleucel (tisa-cel) depending on the availability of manufacturing slots. All patients received a cyclophosphamide and fludarabine-containing lymphodepleting regimen. Response (categorized as complete or partial response) or no response (stable or progressive disease) assessment was done at 1, 3, 6 months and then every 6 months per institutional practice and based on Lugano critera^18^. Primary outcomes were response at 6 months and cumulative incidence of relapse/progression (CIR). Secondary outcomes were progression-free survival (PFS) and overall survival (OS). The study was approved by the review board of Saint-Louis hospital, Assistance Publique-Hôpitaux de Paris (BIOCART-CPP 2019-77) and all patients signed informed consent prior to treatment.

HLA-A, -B, -C, -DRB1, -DQA1, -DQB1 and -DPB1 genotyping was performed using the NGSgo HLA Typing Assay (GENDX, Utrecht, The Netherlands) and sequencing on a Miseq platform (Illumina, San Diego, USA). The corresponding amino acid sequences of the peptide-binding region (encoded by exons 2 and 3 for class 1 genes, and exon 2 for class 2 genes) were extracted from the IMGT/HLA database^19^. The divergence (HED) between the peptide-binding regions of the two HLA alleles at each locus was calculated using the Grantham distance^8^. For HLA-DQA1, the amino acid at position 56 was ignored in the HED calculation because of a deletion mutation in several alleles.

Continuous variables are presented as median and range, categorical variables as number and percentage. Comparison between groups was evaluated by unpaired t-test or Mann-Whitney test for continuous variables, and Fisher’s exact test for categorical variables. CIR was estimated using non-relapse mortality (NRM) as a competing risk and groups were compared with the Gray’s test. PFS and OS were estimated by the Kaplan-Meier method and groups were compared with the log-rank test. Multivariable analyses were performed using logistic regression for response at 6 months, Fine-Gray subdistribution hazard regression for CIR, and Cox proportional hazards model for PFS and OS, including all variables statistically significant (*p* < 0.05) in the univariable analysis. Analysis was conducted using GraphPad Prism 10.0 and R 4.2.1, RStudio, and the tidycmprsk package.

## Results and Discussion

The characteristics of the 127 study patients are shown in Table 1. Among them, 63 (49.6%) were responders (R), defined as those who showed response at 6 months after CAR T-cell infusion, and 64 (50.4%) were non responders (NR) due to disease relapse or death at the 6-month timepoint. Eleven patients died free from relapse after 6 months, including 7 from COVID-19. Median follow-up was 19 months. At day 1000, the estimated CIR was 53.6 %, PFS 38.0 % and OS 48.0 % (**Figure 1A**). At the time of decision to proceed to CAR T-cell therapy, extranodal sites ≥ 2 and high IPI were associated with a higher risk for non-response. At the time of treatment, poor PS, high IPI, elevated LDH levels, high total metabolic tumor volume and the type of CAR T-cell product (tisa-cel versus axi-cel) were associated with an increased risk of treatment failure.

**Table 1.**
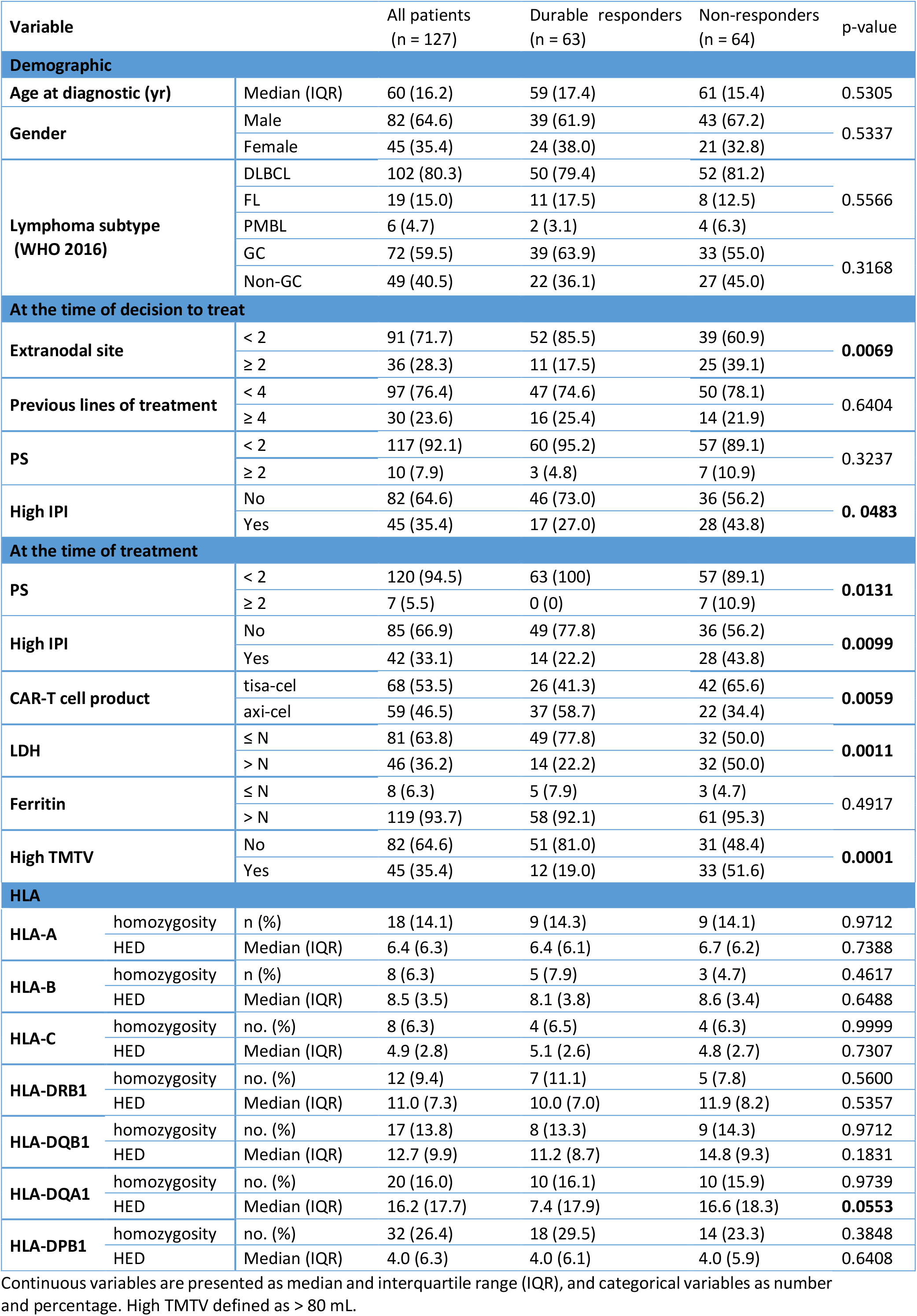
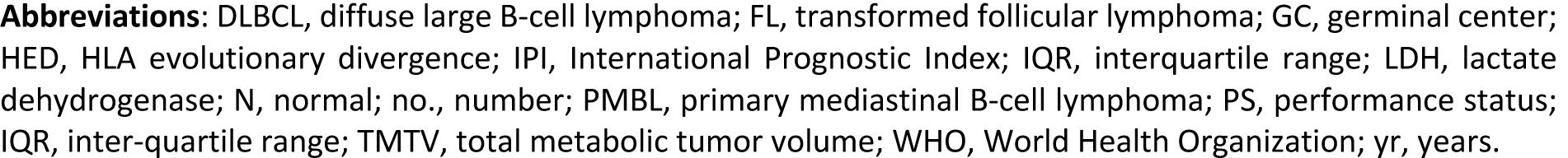
Characteristics of the 127 patients according to the response at 6 months after CART-cell treatment.

**Figure 1.**
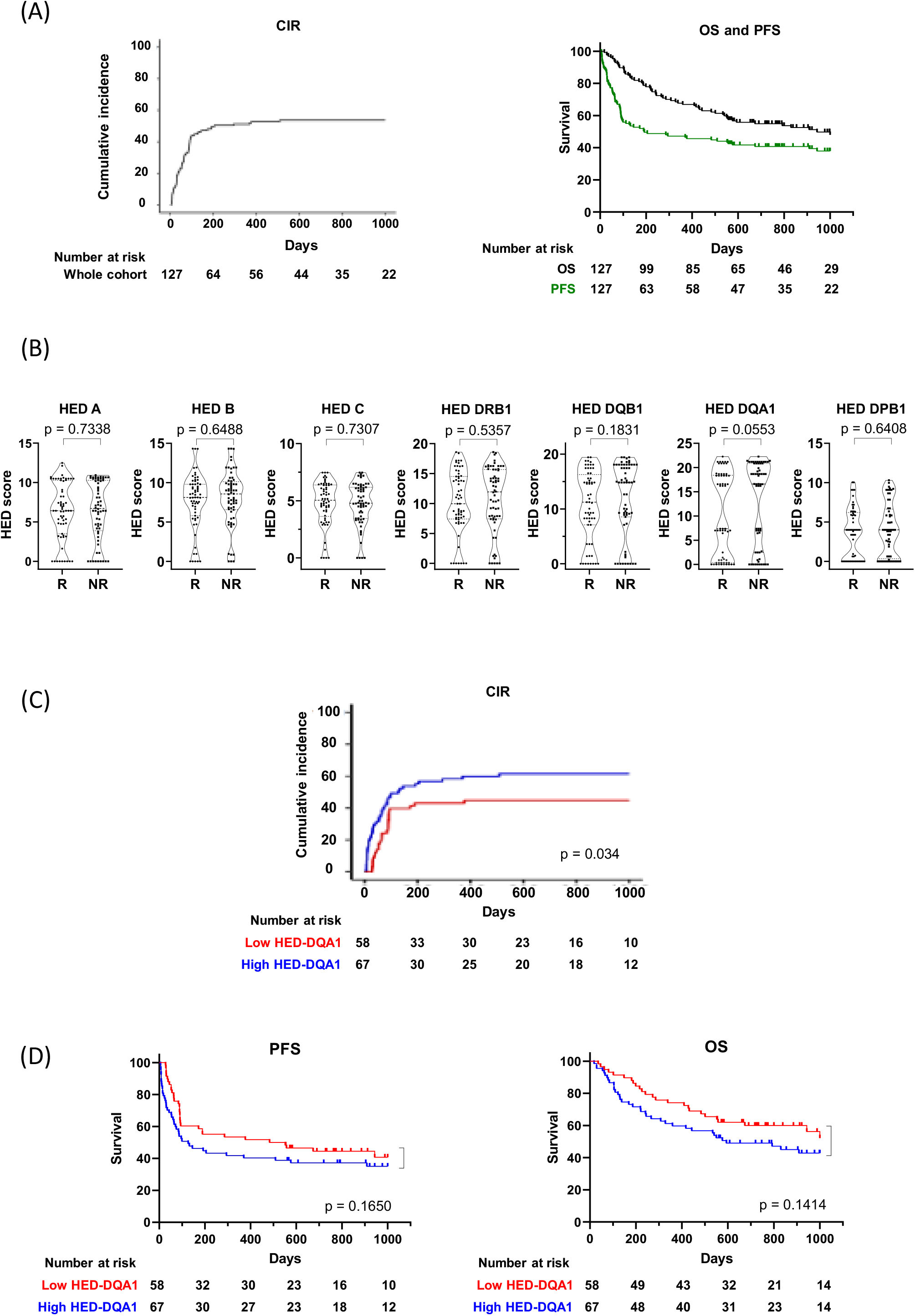
Impact of HED on response to CAR-T cell therapy. **A)** 1000-day estimates of CIR (left panel), PFS and OS (right panel) in the 127 rrLBCL patients treated with commercial CAR-T cells. **B)** Distributions of HED values at the different HLA loci according to response at 6 months. R: durable response (n = 63); NR: non-response (n = 64). **C)** CIR is lower in patients with low HED-DQA1 (ie., below the median in the entire cohort) compared to those with high HED. Gray’s test with NRM as a competing risk. **D)** PFS and OS tend to be increased in patients with low (i.e., below median) HED at the DQA1 locus. *P*-values comparing groups, log-rank test.

We analyzed the potential impact of HLA diversity on the chances of a durable response assessed by the response at 6-months and CIR. There was no effect of HLA heterozygosity. We also did not observe any significant difference in the distribution of HED at any locus between responders and non-responders, except at the HLA-DQA1 locus (Table 1, **Figure 1B**). Intriguingly, HED-DQA1 was lower in responders (median, 7.37 in responders versus 16.6 in non-responders, *p* = 0.055). Moreover, CIR was lower in patients with low HED-DQA1 (ie., below the median in the entire cohort) compared to those with high HED (45% vs 61%, *p* = 0.034) (**Figure 1C)**. Overall, PFS and OS tended to be higher in patients with low HED-DQA1 (**Figure 1D)**.

Upon these unexpected results, we examined the distribution of HED at the HLA-DQA1 locus and observed that HED values were distributed into 3 distinct clusters of very high, low and very low divergence (HED >16, 2.5 <HED <7.5, and HED <1.5, respectively) in contrast to a more continuous distribution at other loci (**Figure 1B)**. When analyzing which pairs of heterozygous DQA1 alleles were represented in these clusters, we found that highly divergent pairs (HED >16) always consisted of an allele of the DQA1*01 supertype together with a non-DQA1*01 allele (i.e., allele of the DQA1*02, 03, 04, 05 or 06 supertype). Conversely, slightly divergent pairs consisted of either two alleles of the DQA1*01 supertype (HED <1.5) or two non-DQA1*01 alleles (HED 2.5-7.5) (**Figure 2A)**.

**Figure 2:**
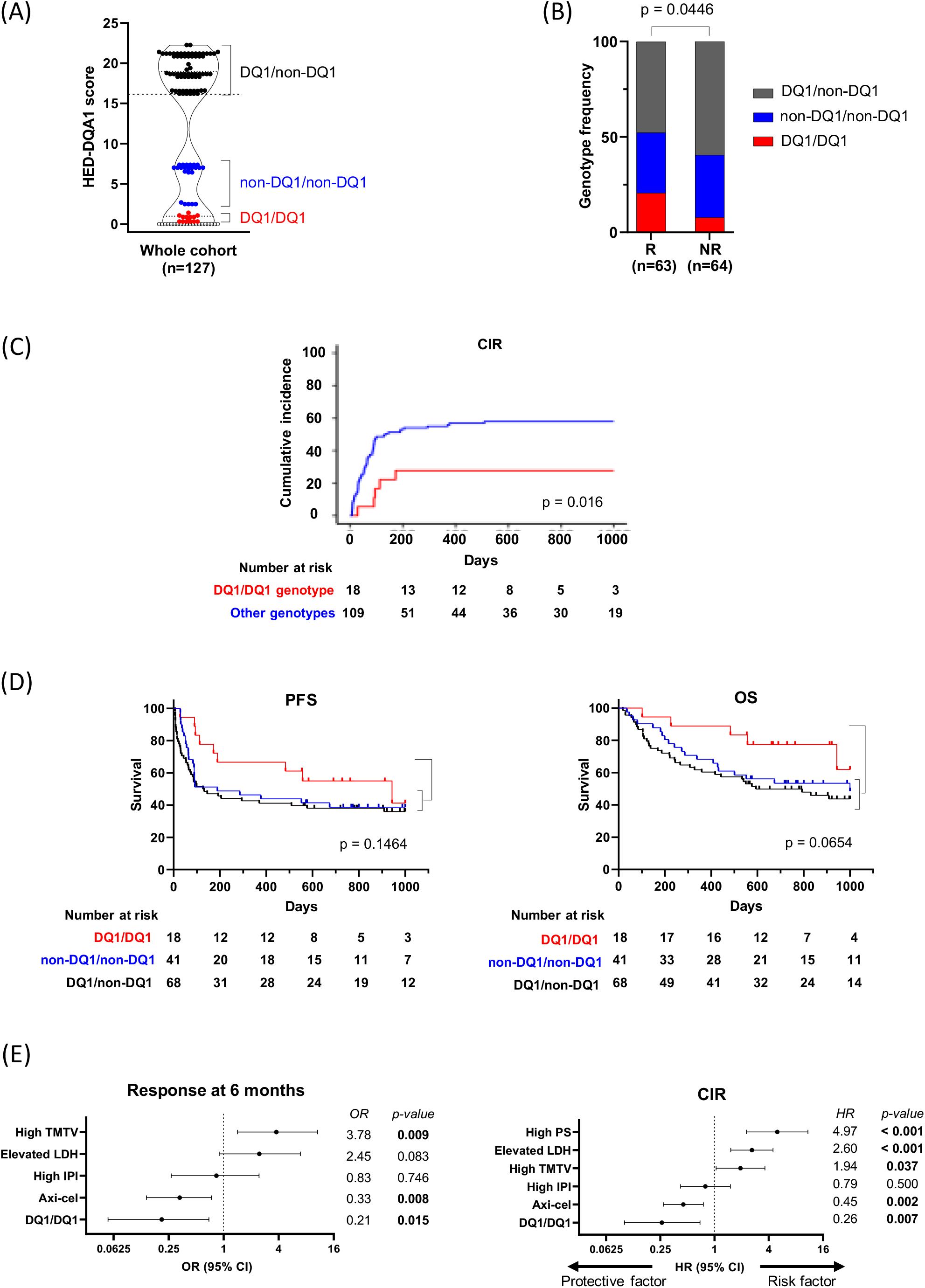
HLA-DQ genotype identifies patients with a better chance of responding to CAR T-cell therapy. **A**) Three clusters of high (>16, black dots), low (2.5-7.5, blue dots) and very low (<1.5, red dots) HED values at the DQA1 locus correspond to distinct heterozygous combinations of DQαβ alleles in the entire patient cohort. HED values equal to zero correspond to homozygotes (empty dots). **B)** Relative distribution of DQ1/DQ1, DQ1/nonDQ1 and nonDQ1/nonDQ1 genotypes according to response at 6 months. R, response; NR, non-response. *P*-value comparing DQ1/DQ1 versus other genotypes, Fisher’s exact test. **C)** The DQ1/DQ1 genotype is associated with lower CIR compared to other genotypes. Gray’s test. **D)** PFS and OS tend to be increased in patients with a DQ1/DQ1 genotype. *P*-values comparing groups, log-rank test. **E)** Multivariable analysis showing the DQ1/DQ1 genotype as an independent protective factor contributing to the response at 6 months and the lower CIR. Forest plots including the respective covariates show adjusted OR (logistic regression) and HR (Fine-Gray regression) with 95% CI.

DQ molecules consist of αβ heterodimers that can be formed as both *cis* and *trans* variants depending on whether the α and β chains are encoded by the DQA1 and DQB1 genes on the same (*cis*) or opposite (*trans*) chromosomes, but not all combinations can form stable DQαβ heterodimers. Interestingly, the HLA-DQA1 and HLA-DQB1 alleles that can pair effectively fall into two mutually exclusive groups hereinafter designed as the DQ1 group (any DQA1*01 allele which can pair with any DQB1*05 or DQB1*06 allele) and the non-DQ1 group (any non-DQA1*01 allele which can pair with any DQB1*02, 03 or 04 allele)^20,21^.

We therefore analyzed the distribution of the DQ1/DQ1, DQ1/non-DQ1 and non-DQ1/non-DQ1 genotypes among patients and found that it significantly differed according to the response (**Figure 2B)**. Patients with a DQ1/DQ1 genotype had a better chance of responding than others (OR = 0.33, 95 % CI 0.12-0.98, *p* = 0.045). Furthermore, the DQ1/DQ1 genotype was associated with a lower incidence of relapse/progression (28 % vs 58 %, p = 0.016) (**Figure 2C)**, and a trend toward higher PFS and OS (**Figure 2D)**. Importantly, after adjusting for other baseline characteristics significantly associated with response, DQ1/DQ1 genotype remained an independent predictor of response (adjusted OR = 0.21, 95 % CI 0.05 -0.69, *p* = 0.0145) and reduced hazard of relapse/progression (adjusted HR = 0.26, CI 0.10 – 0.69, p= 0.007) (**Figure 2E)**.

Altogether, these results in a relatively small cohort of patients point to the HLA-DQ locus as a driver of durable response to CAR-T cell treatment in LBCL patients and suggest a critical role for cellular cross-talk between CAR-T cells and host immune cells in long-term tumor surveillance. Although CAR-T cells act primarily in an HLA-independent fashion, recent studies have highlighted the critical role of CAR T-cell-derived IFNγ in promoting the recruitment of immune cells at the tumor site, upregulating MHC class 2 at the surface of antigen presenting cells and increasing presentation of tumor antigens secondary to antigen spreading^3,4,22,23^. Such mechanisms may overcome tumor heterogeneity or antigen-escape variants and contribute to a durable response after CAR T-cell therapy. The mechanism by which DQ1 and non-DQ1 molecules have opposite effects in this setting remains unclear, but evidence for their distinct function has already been suggested by their opposite impact on the risk of relapse after hematopoietic stem cell transplantation^18^. DQ1 molecules might be more sensitive to IFNγ-induced upregulation or less prone to HLA somatic mutations or allelic losses, thus ensuring sustained presentation of tumor antigens to endogenous T cells. Alternatively, the presence of non-DQ1 molecules (and therefore a higher HLA-DQ diversity) might be associated with a broader repertoire of regulatory T cells in the tumor microenvironment, thus participating in treatment failure. Although our study is limited by the small number of patients, it opens the way for more in-depth investigations on the impact of HLA-DQ genotype on the endogenous anti-tumor response in larger series of CAR-T cell-treated patients.

## Data Availability

All data produced in the present study are available upon reasonable request to the authors

## Acknowledgments

This work was supported by Fondation ARC pour la recherche sur le cancer (*PREDICARTe* ARCTHEM2021010002907).

## Authorship contributions

M.L-M, V.A and S.C-Z conceived and designed the study; M.L-M and J.V performed the experiments; M.L-M and V.A analyzed the data; R.D-B, C.T, and J-L.T contributed with data acquisition; M.L-M, V.A and S.C-Z wrote the manuscript; all authors reviewed the manuscript.

## References

1. Ruella M, Korell F, Porazzi P, Maus MV. Mechanisms of resistance to chimeric antigen receptor-T cells in haematological malignancies. Nat Rev Drug Discov. 2023;22(12):976–995. doi:10.1038/s41573-023-00807-1

2. Shah NN, Fry TJ. Mechanisms of resistance to CAR T cell therapy. Nat Rev Clin Oncol. 2019;16(6):372–385. doi:10.1038/s41571-019-0184-6

3. Boulch M, Cazaux M, Loe-Mie Y, et al. A cross-talk between CAR T cell subsets and the tumor microenvironment is essential for sustained cytotoxic activity. Sci Immunol. 2021;6(57):eabd4344. doi:10.1126/sciimmunol.abd4344

4. Ma L, Hostetler A, Morgan DM, et al. Vaccine-boosted CAR T crosstalk with host immunity to reject tumors with antigen heterogeneity. Cell. 2023;186(15):3148–3165.e20. doi:10.1016/j.cell.2023.06.002

5. Thursz MR, Thomas HC, Greenwood BM, Hill AV. Heterozygote advantage for HLA class-II type in hepatitis B virus infection. Nat Genet. 1997;17(1):11–12. doi:10.1038/ng0997-11

6. Arora J, Pierini F, McLaren PJ, Carrington M, Fellay J, Lenz TL. HLA Heterozygote Advantage against HIV-1 Is Driven by Quantitative and Qualitative Differences in HLA Allele-Specific Peptide Presentation. Mol Biol Evol. 2020;37(3):639–650. doi:10.1093/molbev/msz249

7. Carrington M, Nelson GW, Martin MP, et al. HLA and HIV-1: heterozygote advantage and B*35-Cw*04 disadvantage. Science. 1999;283(5408):1748–1752. doi:10.1126/science.283.5408.1748

8. Pierini F, Lenz TL. Divergent Allele Advantage at Human MHC Genes: Signatures of Past and Ongoing Selection. Mol Biol Evol. 2018;35(9):2145–2158. doi:10.1093/molbev/msy116

9. Chowell D, Morris LGT, Grigg CM, et al. Patient HLA class I genotype influences cancer response to checkpoint blockade immunotherapy. Science. 2018;359(6375):582–587. doi:10.1126/science.aao4572

10. Chowell D, Krishna C, Pierini F, et al. Evolutionary divergence of HLA class I genotype impacts efficacy of cancer immunotherapy. Nat Med. 2019;25(11):1715–1720. doi:10.1038/s41591-019-0639-4

11. Lee CH, DiNatale RG, Chowell D, et al. High Response Rate and Durability Driven by HLA Genetic Diversity in Patients with Kidney Cancer Treated with Lenvatinib and Pembrolizumab. Mol Cancer Res MCR. 2021;19(9):1510–1521. doi:10.1158/1541-7786.MCR-21-0053

12. Roerden M, Nelde A, Heitmann JS, et al. HLA Evolutionary Divergence as a Prognostic Marker for AML Patients Undergoing Allogeneic Stem Cell Transplantation. Cancers. 2020;12(7):1835. doi:10.3390/cancers12071835

13. Merli P, Crivello P, Strocchio L, et al. Human leukocyte antigen evolutionary divergence influences outcomes of paediatric patients and young adults affected by malignant disorders given allogeneic haematopoietic stem cell transplantation from unrelated donors. Br J Haematol. 2023;200(5):622–632. doi:10.1111/bjh.18561

14. Pagliuca S, Gurnari C, Hercus C, et al. Leukemia relapse via genetic immune escape after allogeneic hematopoietic cell transplantation. Nat Commun. 2023;14(1):3153. doi:10.1038/s41467-023-38113-4

15. Féray C, Taupin JL, Sebagh M, et al. Donor HLA Class 1 Evolutionary Divergence Is a Major Predictor of Liver Allograft Rejection : A Retrospective Cohort Study. Ann Intern Med. 2021;174(10):1385–1394. doi:10.7326/M20-7957

16. Wang SS, Carrington M, Berndt SI, et al. HLA Class I and II Diversity Contributes to the Etiologic Heterogeneity of Non-Hodgkin Lymphoma Subtypes. Cancer Res. 2018;78(14):4086–4096. doi:10.1158/0008-5472.CAN-17-2900

17. Wang QL, Wang TM, Deng CM, et al. Association of HLA diversity with the risk of 25 cancers in the UK Biobank. EBioMedicine. 2023;92:104588. doi:10.1016/j.ebiom.2023.104588

18. Cheson BD, Fisher RI, Barrington SF, et al. Recommendations for initial evaluation, staging, and response assessment of Hodgkin and non-Hodgkin lymphoma: the Lugano classification. J Clin Oncol Off J Am Soc Clin Oncol. 2014;32(27):3059–3068. doi:10.1200/JCO.2013.54.8800

19. Robinson J, Halliwell JA, Hayhurst JD, Flicek P, Parham P, Marsh SGE. The IPD and IMGT/HLA database: allele variant databases. Nucleic Acids Res. 2015;43(Database issue):D423–431. doi:10.1093/nar/gku1161

20. Petersdorf EW, Bengtsson M, Horowitz M, et al. HLA-DQ heterodimers in hematopoietic cell transplantation. Blood. 2022;139(20):3009–3017. doi:10.1182/blood.2022015860

21. Liblau RS, Latorre D, Kornum BR, Dauvilliers Y, Mignot EJ. The immunopathogenesis of narcolepsy type 1. Nat Rev Immunol. Published online July 3, 2023. doi:10.1038/s41577-023-00902-9

22. H Kim R, Plesa G, Gladney W, et al. Effect of chimeric antigen receptor (CAR) T cells on clonal expansion of endogenous non-CAR T cells in patients (pts) with advanced solid cancer. Journal of Clinical Oncology. May 2017:3011–3011.

23. Ma L, Dichwalkar T, Chang JYH, et al. Enhanced CAR-T cell activity against solid tumors by vaccine boosting through the chimeric receptor. Science. 2019;365(6449):162–168. doi:10.1126/science.aav8692

